# Frequent, Persistent, and Yearly Inpatient Utilization Across a Multi-Hospital Government Health System in Jeddah, Saudi Arabia: A Retrospective Three-Definition Analysis (2022–2024)

**DOI:** 10.64898/2026.07.08.26357541

**Authors:** Shada Baoum, Rajaa Al-Raddadi, Abdullah Alsahafi, Zaki Algasemi

## Abstract

**Background:** A small proportion of hospitalized patients generates a disproportionate share of inpatient admissions, bed-day utilization, and associated health expenditure globally.^1,2^ In Saudi Arabia, where Vision 2030 mandates measurable reductions in preventable hospitalizations^27^ and hospitals consume approximately 79% of public health expenditure,^23^ population-level evidence on inpatient frequent utilization is absent from the published literature. A key methodological limitation of existing studies is reliance on a single threshold that cannot distinguish acute high-frequency episodes from sustained multi-year hospital dependence.

**Methods:** A retrospective cross-sectional study analyzed electronic health records from three public hospitals in Jeddah — East Jeddah Hospital (EJH), King Abdul-Aziz Hospital (KAAH), and Thagher Hospital (TH) — for January 2022 to December 2024. Records from two clinical information systems (Oasis at KAAH and TH; Careware at EJH) were harmonized using an eight-stage data quality protocol applied to 258,391 raw encounters, yielding a final cohort of 82,160 unique patients and 100,685 valid inpatient visits. Three complementary definitions were applied: Frequent Utilizer (FU: ≥3 admissions within any rolling 365-day window4,6), Persistent Utilizer (PU: ≥3 admissions with ≥24 months between first and last), and Yearly Utilizer (YU: ≥1 admission in each of 2022, 2023, and 2024). Analyses were conducted in JASP 0.95.4 (JASP Team, 2025).^36^

**Results:** FU prevalence was 2.96% (n=2,434), PU 0.60% (n=494), and YU 0.62% (n=507). Overlap analysis identified 177 compound utilizers (0.22%) satisfying all three criteria simultaneously, with a median of 7 admissions and 33.44 bed days — more than thirteen times the standard patient median. Strikingly, compound utilizers had the youngest median age of any utilizer group (24 years), while Saudi nationality concentration rose progressively from 75.0% in standard patients to 87.6% in compound utilizers, and female predominance was highest in the persistence-defined groups (PU-only 62.9%, YU-only 63.6%). All three ANOVA models confirmed significant utilizer status × hospital interactions (all p<.001). Logistic regression confirmed age, Saudi nationality, and hospital as independent predictors across all definitions. A gender discrepancy — significant for males in FU Model 1 (OR=1.090, p=.039) but not Model 2 (p=.181) — was attributable to age confounding.

**Conclusions:** Approximately one in thirty-four inpatients meets the FU criterion in this Jeddah system, with significant between-hospital variation. The three-definition framework reveals clinically distinct utilization phenotypes invisible to any single threshold, including compound utilizers with extraordinary burden and unexpectedly young age, and persistent users entirely missed by annual-window definitions. Saudi nationality is the strongest and most consistent predictor across all definitions. Integrated clinical pathways connecting primary care and community services to hospital care, with shared accountability for quality across levels, are the recommended system response aligned with Vision 2030.

## Background

A globally consistent finding across health systems is the concentration of inpatient resource use among a minority of patients.^1,2^ Studies from the United States, Europe, Australasia, and East Asia demonstrate that between three and ten per cent of hospitalized patients account for the majority of admissions, bed-days, and associated costs — a concentration rooted in the recurrent acute decompensation of inadequately managed chronic non-communicable disease (NCD).^3,5,16^ Understanding who these patients are is a prerequisite for designing effective preventive, primary care, and hospital-based interventions.

Saudi Arabia is a particularly important yet understudied context for this inquiry. Epidemiologically, the Kingdom carries an NCD burden among the highest globally: diabetes prevalence in Jeddah adults aged ≥50 years reaches 46% in men and 44% in women,20 and rising rates of hypertension, obesity, and cardiovascular disease drive a substantial and growing hospitalization burden.^18,21,22,24^ Structurally, approximately 79% of public health expenditure is directed to hospital-based services^25^ — nearly double the OECD average — while primary care remains comparatively underdeveloped in capacity and quality.^19,26,27,28^ The Vision 2030 Health Sector Transformation Programme has set explicit targets for reducing preventable hospitalizations and shifting chronic disease management to primary care,25 but achieving these targets requires a baseline characterization of who is driving inpatient demand — which does not currently exist in the published literature for Saudi Arabia.

A critical and underappreciated methodological limitation of the existing frequent utilizer literature is reliance on a single threshold applied within a fixed window — most commonly three or more admissions within a 12-month period.^4,5,6^ While this captures patients whose admissions cluster intensively within a year, it misses those who accumulate admissions slowly across multiple years without ever meeting the annual threshold, and fails to distinguish patients who had a single difficult year from those with genuinely sustained multi-year hospital dependence — a distinction with direct implications for intervention design.^3,7^

This study introduces three complementary definitions applied simultaneously to the same cohort, enabling the first characterization of the overlap structure between different utilization phenotypes. It is also the first population-level study of all-cause frequent inpatient utilization from a Saudi multi-hospital system. Two contextual factors deserve acknowledgement as potential contributors to the conservative prevalence estimates expected in this system: first, Saudi Arabia’s relatively young population structure^33^ is expected to be healthier on average than the aging Western populations in which most frequent utilizer literature was generated, reducing the absolute burden of chronic multimorbidity that drives recurrent admission; and second, all three study hospitals are general hospitals with active Obstetrics and Gynecology and Pediatrics departments, whose patients — with typically shorter stays and lower readmission rates — dilute both the overall average length of stay and the proportion of patients meeting any frequent utilization threshold compared with specialist medical or surgical facilities.

## Methods

### Study Design and Setting

This was a retrospective cross-sectional study of routinely collected electronic health record (EHR) data from three public Ministry of Health hospitals in Jeddah, Saudi Arabia: East Jeddah Hospital (EJH), King Abdul-Aziz Hospital (KAAH), and Thagher Hospital (TH). EJH is a large secondary care general hospital serving central Jeddah and operated the Careware clinical information system. KAAH is a tertiary general hospital and operated the Oasis system. TH is a secondary care general hospital with a training function and also operated Oasis. The study period was January 1, 2022, to December 31, 2024 — the first complete three-year window of normalized inpatient activity following the COVID-19 pandemic, during which hospitalization patterns across Saudi Ministry of Health facilities had been substantially disrupted by the events of 2020–2021. The study was conducted and reported in accordance with the STROBE guidelines for observational research.^35^

### Data Sources and Harmonization

Source files comprised six annual Oasis extracts from KAAH and TH (2022, 2023, and 2024 for each hospital; date format DD-MM-YYYY with time in a separate column) and one Careware file from EJH spanning January 2022 to August 2025 (date format MM/DD/YYYY HH:MM:SS with time embedded in the date field). Two supplementary files provided bed inventory and test patient identifiers. Raw import yielded 258,391 encounter rows. Table 1 presents the complete data attrition sequence.

**Table 1.** Data attrition sequence from raw import to final analytic cohort.

| Stage | Encounters | Change |
| --- | --- | --- |
| Raw import (all three hospitals) | 258,391 | — |
| After test record removal (EJH; Round 1) | 258,234 | −157 |
| After 2025 admission exclusion (EJH/Careware only) | 249,665 | −8,569 |
| After null discharge date exclusion | 249,136 | −529 |
| After ICD-10 deduplication (unique encounters) | 100,730 | consolidation |
| After negative LOS exclusion | 100,685 | −45 |
| **Final unique patients** | **82,160** | — |

Negative LOS = clinical length of stay < 0 days. ICD-10 deduplication collapsed same-patient same-day multi-code rows into unique encounters; average 2.47 codes per encounter retained.

An eight-stage cleaning and harmonization protocol was applied in DBeaver 25.3.3 (PostgreSQL), with each stage implemented as a SQL view for full auditability30: (i) removal of 157 test encounter records using hospital-provided EJH identifier lists (54 file numbers hardcoded in SQL); (ii) nationality standardization to Saudi / Non-Saudi / Unknown; (iii) gender standardization from M, F, U, Male, Female, and Un-specified to Male, Female, and Unknown; (iv) ISO 8601 timestamp harmonization via SQL regex parsing, reconciling the MM/DD/YYYY HH:MM:SS Careware format with the DD-MM-YYYY Oasis format, combining the date and separate time columns for KAAH and TH records; (v) age standardization from date-of-birth strings, integers, and compound year-month-day formats (e.g., ‘02Y 03M 10D’) to decimal years, with biologically impossible negative values (n=112) and extreme outliers exceeding 110 years (n=110), consistent with administrative placeholder dates of 01/01/1900, set to missing; (vi) exclusion of 8,569 admissions dated on or after January 1, 2025, all of which originated from the EJH Careware file, which extended beyond the study window; (vii) exclusion of 529 encounters within the 2022–2024 window with null discharge dates, representing patients whose records were never formally closed in the system — these encounters could not contribute to length-of-stay or bed-day calculations; (viii) ICD-10 deduplication consolidating same-patient same-day multi-code encounter rows into unique clinical encounters with a diagnosis count column. Deduplication was performed by grouping on the composite key of patient identifier, hospital, and exact admission timestamp, ensuring that only records sharing an identical admission timestamp — representing the same clinical visit entered as multiple diagnostic code rows — were consolidated. This composite key approach was developed for this study to address the cross-system file-number non-uniqueness identified during data governance review.

A critical data governance finding was that patient file numbers are not unique across hospitals — the same number can identify different patients at different facilities. All patient-level operations used composite keys combining file number and hospital identifier throughout the entire pipeline, including the utilizer classification views and all join operations.

The eight-stage protocol yielded a final analytic encounter base of 100,685 valid inpatient visits. Patient-level records were derived from this encounter base by aggregating on the composite key of patient identifier and hospital, retaining the first admission as the index visit and calculating utilization metrics — total admission count, cumulative bed days, and age at index visit — across all encounters attributed to each unique patient. This aggregation produced the final cohort of 82,160 unique patients on which all subsequent analyses were conducted.

Sub-24-hour visits (n=20,227) were retained following inspection of their age group and hospital distribution, which confirmed wide representation across all age categories and all three facilities, consistent with genuine clinical encounters including newborn deliveries, day procedures, and brief observation admissions. Planned recurring procedures such as scheduled dialysis or blood transfusions that generate sub-24-hour admission records are intentionally included, as they represent genuine inpatient resource consumption and contribute directly to the burden profile of patients with chronic conditions. Their inclusion is consistent with the all-cause, system-level scope of this study; exclusion would undercount the true inpatient footprint of the highest-burden patients, particularly those with renal disease or haematological conditions.

A total of 115 patients (0.14%) had missing age at index visit after cleaning. These were concentrated at EJH in 2022 (n=43) and 2023 (n=64), with no missing age at EJH in 2024, suggesting a time-limited data entry issue at EJH that resolved by 2024. These patients were excluded from age-based analyses only.

### Utilizer Definitions

Three classification views were constructed on the clean visit base (vw_valid_visits) using composite patient keys. Frequent Utilizer (FU): ≥3 admissions within any rolling 365-day window.^4,6,7,8,9,10^ Although the rolling-window approach originates in emergency department frequent attender literature,6,7 it is methodologically portable to inpatient admission data and avoids the artificial deflation of prevalence caused by calendar-year boundaries. A self-join approach anchored each admission as a reference point and counted distinct admission dates within the following 365 days, with a DISTINCT outer query preventing double-counting across overlapping windows. For transparency, an earlier iteration applied a study-period definition (≥3 admissions over the full 36-month window), which yielded 3,255 patients (3.96%); the rolling-window definition is methodologically more rigorous and excludes patients whose admissions were spread too thinly to constitute a high-frequency episode within any 12-month period. Persistent Utilizer (PU): ≥3 admissions with ≥24 months between first and last admission. Captures long-horizon hospital dependence regardless of annual clustering. Yearly Utilizer (YU): ≥1 admission in each of the three calendar years 2022, 2023, and 2024. Captures sustained system engagement across the entire observation window. The three definitions are mutually overlapping; patients satisfying all three criteria are referred to as compound utilizers — a term introduced by the present authors to describe this intersection group, following the precedent of overlap-based high utilizer characterization in the literature.^15^

The ≥3-admission threshold for FU classification is consistent with the closest published inpatient precedents: Low et al. defined frequent inpatient admission as three or more hospitalizations within 12 months in a Singapore government hospital cohort^8^; Longman et al. applied the same threshold to older Australians with chronic ambulatory care sensitive conditions^9^; and the CHAMP randomized controlled trial used three or more inpatient readmissions in the prior year as its enrolment criterion.^10^ The rolling 365-day window applied here is methodologically more rigorous than a fixed calendar-year window, capturing admissions that cluster within any consecutive 12-month period regardless of where that period falls in the study window and thereby avoiding the artificial deflation of prevalence that occurs when admissions straddle a calendar-year boundary.

### Statistical Analysis

Analyses were performed in JASP 0.95.4 (Apple Silicon; JASP Team, 2025).^36^ Age was compared using the Mann-Whitney U test (rank-biserial correlation; pre-specified primary test given right-skewed utilization data) and Student’s independent samples t-test (Cohen’s d). Categorical variables were compared using Pearson’s chi-squared with Cramér’s V. Two-way ANOVA examined total bed-day utilization as a function of utilizer status, hospital, and their interaction. Two binary logistic regression models were fitted per definition: Model 1 with continuous age, hospital, nationality, and gender; Model 2 with a six-level age group factor (Infant <1, Child 1–12, Adolescent 13–17 [reference], Young Adult 18–44, Middle Age 45–64, Senior ≥65) replacing continuous age. Model fit was assessed by Nagelkerke R^2^.^37^ Unknown gender (n≤182 across groups) produced computationally extreme OR values from near-zero cell counts and is excluded from interpretation. Two-tailed α=0.05. Artificial intelligence tools were used in the preparation of this manuscript for linguistic refinement, structural organisation, and iterative drafting. All data collection, statistical analysis, interpretation of results, and conclusions are the sole responsibility of the authors.

### Ethics

This study was approved by the Institutional Review Board — Jeddah (Local IRB, Jeddah Health Affairs; KACST registration number H-02-J-002; IRB approval number A02254; initial approval 16 July 2025; final approval 18 May 2026). The study was conducted in accordance with the Declaration of Helsinki and KACST Good Clinical Practice regulations.

### Consent to Participate

Individual informed consent was not required and was waived by the Institutional Review Board — Jeddah. The study used routinely collected administrative electronic health record data provided by each participating hospital. Source records contained administrative patient identifiers used solely for within-pipeline record linkage and deduplication; these identifiers were not used in any analysis and no individually identifiable data were retained in the final analytic dataset. No contact was made with patients or their representatives at any stage of the study.

## Results

### Study Cohort and Utilizer Overlap

The final cohort comprised 82,160 unique patients with 100,685 valid inpatient visits across the three study hospitals. Table 2 presents the complete overlap structure of the three definitions with demographic profiles and resource utilization for each group.

**Table 2.** Overlap between Frequent (FU), Persistent (PU), and Yearly (YU) Utilizer definitions: demographics and bed-day utilization across all groups (N=82,160). ★ = youngest median age among all utilizer groups. † = entirely invisible under annual-window FU definition.

| Group | n | % total | Saudi % | Female % | Median age (IQR) | Median visits (IQR) | Median bed days (IQR) | Note |
| --- | --- | --- | --- | --- | --- | --- | --- | --- |
| **FU+PU+YU (compound)** | **177** | **0.22** | **87.6** | **53.7** | **24 (31)** | **7 (6)** | **33.44 (47.48)** | **★** |
| FU+PU | 55 | 0.07 | 85.5 | 60.0 | 30 (38) | 5 (1) | 22.57 (22.29) |  |
| FU+YU | 112 | 0.14 | 83.9 | 58.0 | 35 (43) | 5 (3) | 19.76 (25.23) |  |
| FU only | 2,090 | 2.54 | 78.7 | 52.2 | 38 (35) | 3 (1) | 12.84 (19.27) |  |
| PU+YU | 119 | 0.14 | 89.1 | 59.7 | 35 (27) | 3 (1) | 10.00 (15.04) |  |
| PU only | 143 | 0.17 | 86.0 | 62.9 | 34 (24) | 3 (0) | 10.27 (11.88) | † |
| YU only | 99 | 0.12 | 88.9 | 63.6 | 30 (45) | 3 (0) | 10.06 (11.46) |  |
| **Standard (none)** | **79,365** | **96.60** | **75.0** | **52.9** | **28 (37)** | **1 (0)** | **2.50 (4.02)** |  |

FU = ≥3 admissions in any rolling 365-day window. PU = ≥3 admissions with ≥24 months between first and last. YU = ≥1 admission each calendar year 2022–2024. Bed days = total economic length of stay. Verification: FU=2,434 ✓| PU=494 ✓| YU=507 ✓| Total=82,160 ✓Three findings from Table 2 warrant immediate attention. First, the 177 compound utilizers had the youngest median age (24 years) of any utilizer group yet consumed a median of 33.44 bed days — more than thirteen times the standard patient. Second, Saudi nationality concentration increased progressively with utilizer complexity, from 75.0% in standard patients to 87.6% in compound utilizers, with PU-only and YU-only groups exceeding 86%. Third, female predominance was substantially higher in persistence-defined groups — PU-only 62.9%, YU-only 63.6% — compared with 52.2% in the FU-only group, suggesting that the chronic multi-year engagement dimension of hospitalization is more female-predominant than the acute clustering dimension.

### Frequent Utilizer Demographics and Bed-Day Burden

2,434 patients (2.96%) were classified as Frequent Utilizers. Intra-hospital FU prevalence was 3.54% at KAAH (n=1,054/29,733), 2.85% at EJH (n=1,202/42,109), and 1.73% at TH (n=178/10,318). Table 3 presents the full demographic comparison.

**Table 3.** Demographic characteristics and bed-day utilization by Frequent Utilizer status (N=82,160).

| Variable | FU (n=2,434) | Non-FU (n=79,726) | Test statistic | p | Effect size |
| --- | --- | --- | --- | --- | --- |
| **Age (years) — mean (SD)** | 38.42 (24.04) | 28.03 (23.06) | t(82,043)=21.88 | < .001 | d=0.450 |
| Age — median (IQR) | 37 (37) | 28 (37) | U=1.218×10<sup>8</sup> | < .001 | r=−0.257 |
| **Bed days — mean (SD)** | 24.49 (31.53) | 5.91 (16.50) | F(1)=1,056.58 | < .001 | η<sup>2</sup>=0.013 |
| Bed days — median (IQR) | 14.34 (22.11) | 2.50 (4.02) |  |  |  |
| **Saudi nationality, n (%)** | 1,940 (79.7%) | 59,700 (75.1%) | χ<sup>2</sup>(2)=32.13 | < .001 | V=0.020 |
| Non-Saudi, n (%) | 483 (19.8%) | 18,828 (23.7%) |  |  |  |
| **Female, n (%)** | 1,283 (52.7%) | 42,276 (53.0%) | χ<sup>2</sup>(2)=3.86 | p=.145 ns | V=0.007 |
| Male, n (%) | 1,150 (47.2%) | 37,269 (46.7%) |  |  |  |
| **Hospital — EJH, n [intra-%]** | 1,202 (49.4%) [2.85%] | 40,907 (51.3%) | χ<sup>2</sup>(2)=91.74 | < .001 | V=0.033 |
| Hospital — KAAH, n [intra-%] | 1,054 (43.3%) [3.54%] | 28,679 (36.0%) |  |  |  |
| Hospital — TH, n [intra-%] | 178 (7.3%) [1.73%] | 10,140 (12.7%) |  |  |  |

Non-FU age: Missing=115 (0.14%), concentrated at EJH in 2022 (n=43) and 2023 (n=64), consistent with a time-limited data entry protocol gap. Intra-hospital FU% shown in brackets. ANOVA for bed days: hospital × FU status interaction F(2,82,154)=27.79, p<.001.

FUs were significantly older (median 37 vs 28 years; U=1.218×10^8^, p<.001, rank-biserial r=™0.257; Cohen’s d=0.450), more likely to be Saudi nationals (79.7% vs 75.1%; χ^2^(2)=32.13, N=81,896, p<.001, V=0.020), and unequally distributed across hospitals (χ^2^(2)=91.74, p<.001, V=0.033). Gender was not a significant bivariate predictor (χ^2^(2)=3.86, p=.145). The two-way ANOVA confirmed highly significant main effects of FU status on bed-day utilization (F(1,82,154)=1,056.58, p<.001, η^2^=0.013) and a significant hospital × FU status interaction (F(2,82,154)=27.79, p<.001). Non-FU mean bed days were 7.39 at KAAH, 5.48 at EJH, and 3.44 at TH — confirming a more complex overall case mix at the tertiary facility that extends beyond its FU population, consistent with its role as a referral center.

### PU and YU Burden

PU and YU ANOVA models similarly showed significant main effects of utilizer status (PU: F(1,82,154)=258.88, p<.001, η^2^=0.003; YU: F(1,82,154)=285.66, p<.001, η^2^=0.003) and significant hospital × utilizer status interactions (PU: F(2,82,154)=16.99, p<.001; YU: F(2,82,154)=16.05, p<.001). PU mean bed days were 29.42 vs 6.32 for non-PU patients; YU mean bed days were 29.85 vs 6.31 for non-YU patients.

### Logistic Regression: FU Predictors

Table 4 presents both FU models. In Model 1, each additional year of age independently associated with 1.9% higher FU odds (OR=1.019, p<.001); Saudi nationality conferred 53.3% higher odds (OR=1.533, p<.001); and male gender was nominally significant (OR=1.090, p=.039). In Model 2, the gender effect attenuated to non-significance (OR=1.059, p=.181), confirming age confounding in the linear specification rather than a genuine independent gender effect — consistent with sex-difference findings in multimorbid frequent utilizers observed in ED-based international cohorts.^34^ The Wald statistic for gender in Model 1 was 4.3; while nominally significant at this sample size, the effect size was negligible and the association was not replicated in the age-group model, indicating that the Model 1 gender coefficient likely reflects residual age confounding rather than an independent gender effect. Senior patients (≥65 years) were borderline non-significant relative to Adolescents (OR=1.201, p=.056).

**Table 4.**
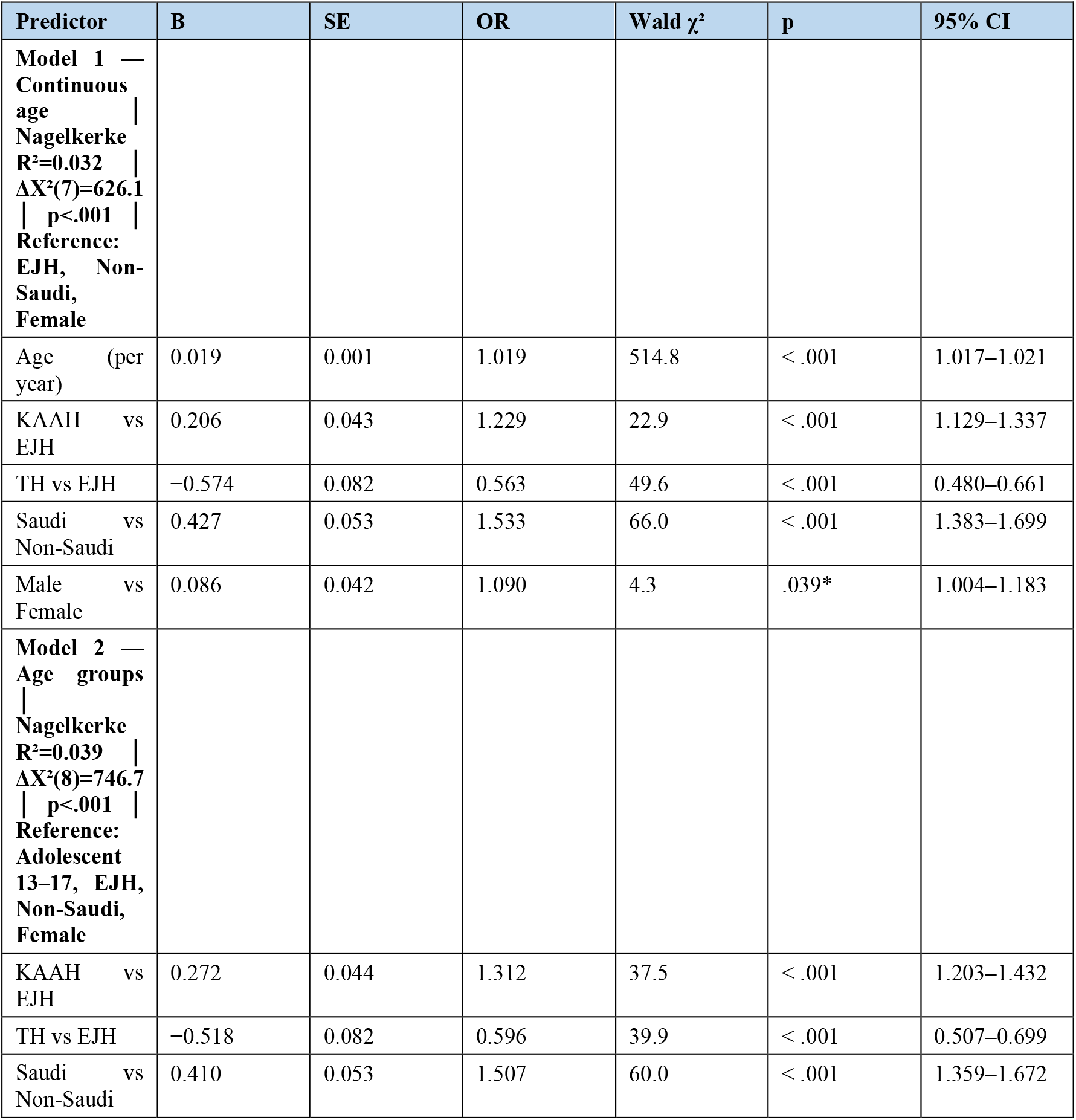
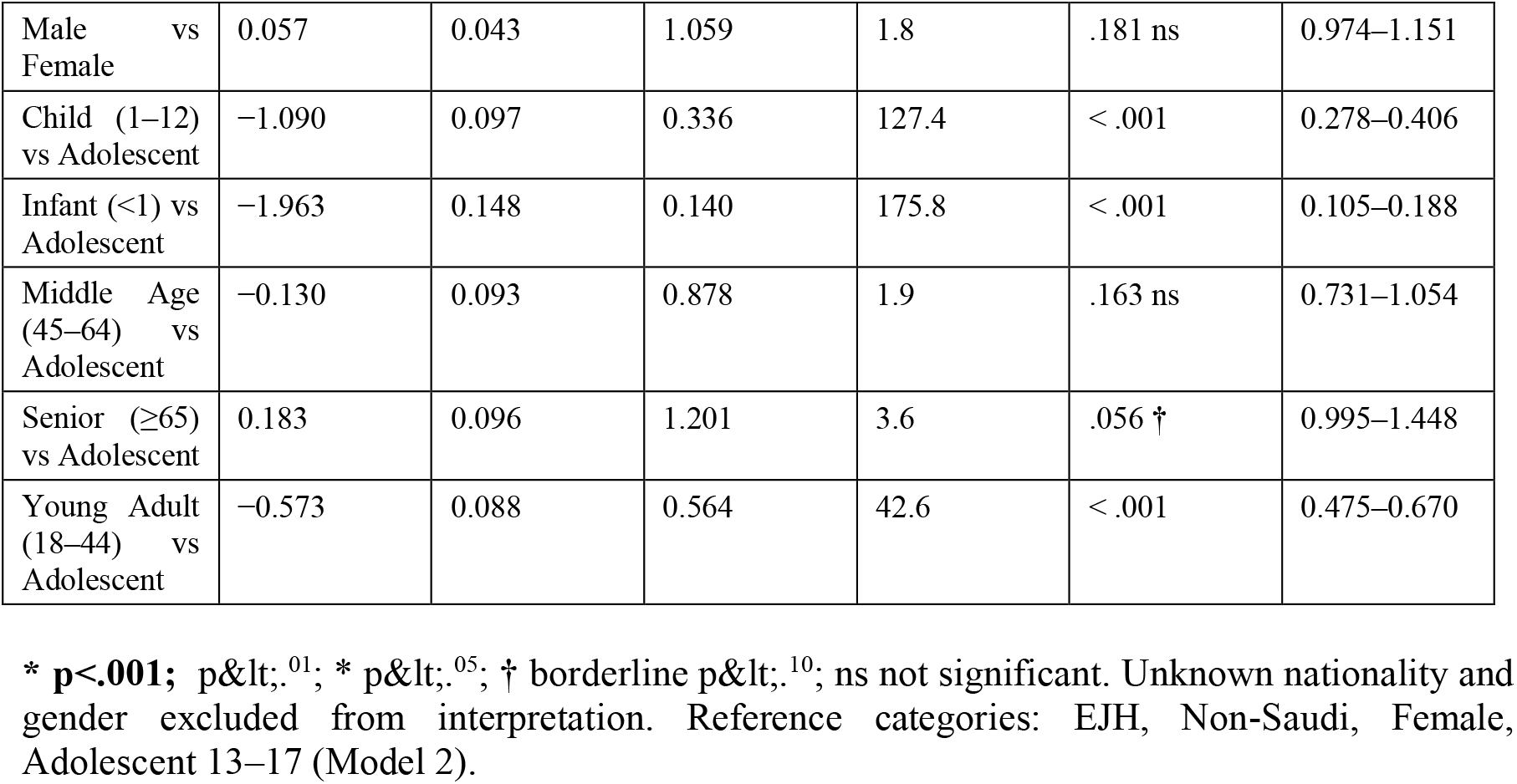
Binary logistic regression: predictors of Frequent Utilizer (FU) status — Model 1 (continuous age) and Model 2 (categorical age groups).

| Predictor | B | SE | OR | Wald $\chi^2$ | p | 95% CI |
| --- | --- | --- | --- | --- | --- | --- |
| **Model 1 —**<br>**Continuous**<br>**age**<br>**Nagelkerke**<br>**R<sup>2</sup>=0.032**<br>**$\Delta X^2(7)=626.1$**<br>**p<.001**<br>**Reference:**<br>**EJH, Non-**<br>**Saudi,**<br>**Female** |  |  |  |  |  |  |
| Age (per<br>year) | 0.019 | 0.001 | 1.019 | 514.8 | < .001 | 1.017–1.021 |
| KA AH vs<br>EJH | 0.206 | 0.043 | 1.229 | 22.9 | < .001 | 1.129–1.337 |
| TH vs EJH | −0.574 | 0.082 | 0.563 | 49.6 | < .001 | 0.480–0.661 |
| Saudi vs<br>Non-Saudi | 0.427 | 0.053 | 1.533 | 66.0 | < .001 | 1.383–1.699 |
| Male vs<br>Female | 0.086 | 0.042 | 1.090 | 4.3 | .039* | 1.004–1.183 |
| **Model 2 —**<br>**Age groups**<br>**Nagelkerke**<br>**R<sup>2</sup>=0.039**<br>**$\Delta X^2(8)=746.7$**<br>**p<.001**<br>**Reference:**<br>**Adolescent**<br>**13–17, EJH,**<br>**Non-Saudi,**<br>**Female** |  |  |  |  |  |  |
| KA AH vs<br>EJH | 0.272 | 0.044 | 1.312 | 37.5 | < .001 | 1.203–1.432 |
| TH vs EJH | −0.518 | 0.082 | 0.596 | 39.9 | < .001 | 0.507–0.699 |
| Saudi vs<br>Non-Saudi | 0.410 | 0.053 | 1.507 | 60.0 | < .001 | 1.359–1.672 |
| Male vs Female | 0.057 | 0.043 | 1.059 | 1.8 | .181 ns | 0.974–1.151 |
| Child (1–12) vs Adolescent | −1.090 | 0.097 | 0.336 | 127.4 | < .001 | 0.278–0.406 |
| Infant (<1) vs Adolescent | −1.963 | 0.148 | 0.140 | 175.8 | < .001 | 0.105–0.188 |
| Middle Age (45–64) vs Adolescent | −0.130 | 0.093 | 0.878 | 1.9 | .163 ns | 0.731–1.054 |
| Senior (≥65) vs Adolescent | 0.183 | 0.096 | 1.201 | 3.6 | .056 † | 0.995–1.448 |
| Young Adult (18–44) vs Adolescent | −0.573 | 0.088 | 0.564 | 42.6 | < .001 | 0.475–0.670 |

\* $p<.001$ ; $p<.01$ ; \* $p<.05$ ; † borderline $p<.10$ ; ns not significant. Unknown nationality and gender excluded from interpretation. Reference categories: EJH, Non-Saudi, Female, Adolescent 13–17 (Model 2).

The Nagelkerke R^2^ values across all six models ranged from 0.020 to 0.039. This modest explanatory power is expected for logistic regression applied to population-level administrative health data, where the binary outcome is rare and is shaped by a large number of unmeasured factors — including primary diagnosis, comorbidity burden, social support, and access to primary care — that are absent from administrative records. The value of these models lies in the direction, magnitude, and cross-model consistency of the OR estimates, which replicate across all six models and are each grounded in a cohort of 82,160 patients.

### Logistic Regression: PU and YU Predictors

Table 5 presents all PU and YU models. Saudi nationality showed substantially stronger effects in PU and YU models than in FU models: PU Model 1 OR=2.486, YU Model 1 OR=2.492, compared with FU Model 1 OR=1.533. A counterintuitive finding emerged in both PU and YU Model 2: Adolescents (the reference group) had higher persistent-utilization odds than all other age groups including Seniors. In PU Model 2, Senior OR=0.657 (p=.031); in YU Model 2, Senior OR=0.711 (p=.062). This contrasts with FU Model 2, where Seniors had the highest odds of any age group (OR=1.201, p=.056). A definition-specific gender finding appeared in YU Model 2: males had significantly lower YU odds than females (OR=0.831, p=.048), consistent with the higher female proportions in persistence-defined groups observed in Table 2.

**Table 5.** Binary logistic regression: predictors of Persistent Utilizer (PU) and Yearly Utilizer (YU) status — Models 1 and 2 for each definition.

| Predictor | B | SE | OR | Wald $\chi^2$ | p | 95% CI |
| --- | --- | --- | --- | --- | --- | --- |
| **PU Model 1**<br>—<br>**Continuous age**<br>**R<sup>2</sup>=0.020**<br>**$\Delta X^2=118.9$**<br>**$p<.001$**<br>**Reference:**<br>**EJH, Non-Saudi, Female** |  |  |  |  |  |  |
| Age (per year) | 0.013 | 0.002 | 1.013 | 47.9 | < .001 | 1.009–1.017 |
| KAAH vs EJH | 0.216 | 0.094 | 1.241 | 5.3 | .021* | 1.033–1.492 |
| TH vs EJH | −0.634 | 0.184 | 0.530 | 11.9 | < .001 | 0.370–0.760 |
| Saudi vs Non-Saudi | 0.911 | 0.139 | 2.486 | 43.2 | < .001 | 1.895–3.263 |
| Male vs Female | −0.133 | 0.092 | 0.875 | 2.1 | .149 ns | 0.731–1.049 |
| **PU Model 2** |  |  |  |  |  |  |
| —<br>**Age groups**<br>**R<sup>2</sup>=0.033**<br>**ΔX<sup>2</sup>=193.5**<br>**p<.001**<br>**Reference:**<br>**Adolescent,**<br>**EJH, Non-**<br>**Saudi,**<br>**Female** |  |  |  |  |  |  |
| KAAH vs EJH | 0.307 | 0.096 | 1.360 | 10.2 | .001** | 1.126–1.643 |
| TH vs EJH | −0.568 | 0.185 | 0.567 | 9.4 | .002** | 0.395–0.814 |
| Saudi vs Non-Saudi | 0.895 | 0.139 | 2.447 | 41.3 | < .001 | 1.862–3.215 |
| Male vs Female | −0.126 | 0.095 | 0.882 | 1.8 | .184 ns | 0.732–1.062 |
| Child (1–12) vs Adolescent | −1.353 | 0.186 | 0.258 | 52.8 | < .001 | 0.179–0.372 |
| Infant (<1) vs Adolescent | −2.608 | 0.352 | 0.074 | 55.0 | < .001 | 0.037–0.147 |
| Middle Age (45–64) vs Adolescent | −0.721 | 0.188 | 0.486 | 14.7 | < .001 | 0.336–0.703 |
| Senior (≥65) vs Adolescent | −0.421 | 0.195 | 0.657 | 4.6 | .031* | 0.448–0.963 |
| Young Adult (18–44) vs Adolescent | −0.768 | 0.164 | 0.464 | 22.0 | < .001 | 0.337–0.639 |
| **YU Model 1**<br>—<br>**Continuous age**<br>**R<sup>2</sup>=0.021**<br>**ΔX<sup>2</sup>=126.0**<br>**p<.001**<br>**Reference:**<br>**EJH, Non-**<br>**Saudi,**<br>**Female** |  |  |  |  |  |  |
| Age (per year) | 0.013 | 0.002 | 1.013 | 52.8 | < .001 | 1.010–1.017 |
| KAAH vs EJH | 0.204 | 0.093 | 1.226 | 4.8 | .028* | 1.022–1.470 |
| TH vs EJH | −0.639 | 0.181 | 0.528 | 12.5 | < .001 | 0.370–0.753 |
| Saudi vs Non-Saudi | 0.913 | 0.136 | 2.492 | 44.8 | < .001 | 1.907–3.256 |
| Male vs Female | −0.110 | 0.091 | 0.896 | 1.5 | .228 ns | 0.750–1.071 |
| **YU Model 2**<br>—<br>**Age groups**<br>**R<sup>2</sup>=0.033**<br>**ΔX<sup>2</sup>=197.1**<br>**p<.001** |  |  |  |  |  |  |
| **Reference:**<br>**Adolescent,**<br>**EJH, Non-**<br>**Saudi,**<br>**Female** |  |  |  |  |  |  |
| KAAH vs<br>EJH | 0.325 | 0.096 | 1.384 | 11.5 | < .001 | 1.147–1.669 |
| TH vs EJH | −0.554 | 0.182 | 0.575 | 9.2 | .002** | 0.402–0.822 |
| Saudi vs<br>Non-Saudi | 0.861 | 0.137 | 2.365 | 39.4 | < .001 | 1.808–3.095 |
| Male vs<br>Female | −0.185 | 0.093 | 0.831 | 3.9 | .048* | 0.692–0.998 |
| Child (1–12)<br>vs<br>Adolescent | −1.240 | 0.174 | 0.289 | 50.9 | < .001 | 0.206–0.407 |
| Infant (<1) vs<br>Adolescent | −2.385 | 0.305 | 0.092 | 61.2 | < .001 | 0.051–0.167 |
| Middle Age<br>(45–64) vs<br>Adolescent | −0.717 | 0.179 | 0.488 | 16.1 | < .001 | 0.344–0.693 |
| Senior (≥65)<br>vs<br>Adolescent | −0.341 | 0.183 | 0.711 | 3.5 | .062 † | 0.497–1.017 |
| Young Adult<br>(18–44) vs<br>Adolescent | −1.066 | 0.160 | 0.344 | 44.3 | < .001 | 0.251–0.471 |

\* $p < .001$ ; $p < .^{.01}$ ; \* $p < .^{.05}$ ; † borderline $p < .^{.10}$ ; ns not significant. Unknown gender excluded. Reference categories: EJH, Non-Saudi, Female, Adolescent 13–17 (Models 2).

## Discussion

### Principal Findings

This study provides the first population-level characterization of frequent inpatient utilization from a Saudi multi-hospital system, applying three complementary definitions simultaneously to reveal a spectrum of recurrent inpatient need invisible to any single threshold. Four findings stand out.

First, compound utilizers — the 177 patients satisfying all three criteria — had a median age of only 24 years yet consumed a median of 33.44 bed days, making them simultaneously the youngest and the highest-burden group in the cohort. This directly challenges the assumption, common in the frequent utilizer literature — including ED-based cohorts^5^ and inpatient studies^16,17^ — that the highest-resource patients are elderly multimorbid patients with inadequately managed chronic disease. In the Jeddah context, we hypothesize that young patients with persistent multi-year inpatient dependence represent a clinically distinct phenotype — most plausibly individuals with chronic high-burden conditions present from early life (e.g., hematologic or renal disorders) — whose inpatient engagement is not discretionary but structurally mandated by a care model that has no viable community-based alternative. This hypothesis is speculative; without ICD-10 diagnostic data linkage it cannot be confirmed. It is, however, a testable and precise prediction for future studies and a specific target for integrated pathway investment.

Second, the 143 PU-only patients — carrying a median of 10.27 bed days, nearly four times the standard patient — would have been entirely invisible under a standard annual-window FU definition. These are individuals with genuine, sustained multi-year hospital dependence who never clustered three admissions within a 12-month period. The PU-only group is the clearest demonstration of the methodological value of the multi-definition framework: single-threshold studies systematically miss this population entirely.

Third, Saudi nationality concentration rose progressively with utilizer complexity, from 75.0% in standard patients to 87.6% in compound utilizers. The substantially stronger Saudi nationality ORs in PU and YU models (both approximately 2.49) compared with the FU model (OR=1.53) suggest that structural access and NCD burden differentials between Saudi nationals and expatriates are amplified among the most persistent hospital users — those whose hospital engagement has become a sustained feature of their healthcare across multiple years.

Fourth, female predominance was substantially higher in persistence-defined groups (PU-only 62.9%, YU-only 63.6%) than in the acute cluster group (FU-only 52.2%), and male gender was a significant protective factor in YU Model 2 (OR=0.831, p=.048). This pattern suggests a specific hypothesis: women in this system may be more likely than men to maintain sustained year-on-year hospital engagement, potentially reflecting higher rates of gynecological, autoimmune, or metabolic conditions requiring repeated planned admission across years. This is a testable prediction for future diagnostic data linkage studies.

### Prevalence in Regional and International Context

The 2.96% FU prevalence is consistent with published inpatient benchmarks. Low et al. found 17.1% of patients in a Singapore government hospital had ≥3 admissions within 12 months^8^ — a higher figure than ours, likely reflecting their broader all-cause definition applied to a higher-acuity tertiary facility. Longman et al. reported a comparable threshold among older Australians with chronic conditions.^9^ The 2.9% reported by Berry et al. in US children’s hospitals using a rolling 365-day window^17^ is the most directly comparable figure. Our 2.96% falls within the 3–8% range reported across international inpatient cohort studies. Two system-level factors likely contribute to the modest Saudi prevalence beyond definitional differences. First, the Saudi population is substantially younger than the aging Western populations in which most frequent utilizer literature was generated: the median age of the entire study cohort was under 30 years, and a younger population structure predicts lower rates of the multimorbidity-driven chronic disease burden that generates the highest rates of frequent utilization.^33^ Second, all three study hospitals are general hospitals with active Obstetrics and Gynecology and Pediatrics departments, whose patients — with typically shorter stays, lower readmission rates, and predominantly acute rather than chronic care needs — dilute both the average length of stay and the proportion of patients meeting any frequent utilization threshold compared with pure medical or specialist facilities. No published inpatient frequent utilizer study exists from Saudi Arabia or the broader GCC region, making direct regional benchmarking impossible. The only available GCC comparator is an ED-based study from Bahrain,^14^ which found a high utilizer prevalence of 3.9% using a four-visit annual threshold. While not directly comparable — ED frequent attenders and inpatient frequent admitters are clinically and operationally distinct cohorts — the Bahraini finding is consistent with the broader pattern of hospital-centered care-seeking in GCC government health systems, where free access without financial deterrents and limited primary care alternatives drive high acute facility utilization across all service types.^23,26,27^

### Saudi Nationality and Structural Determinants

Saudi nationality was the strongest and most consistent independent predictor across all six regression models. Three structural factors may contribute to this finding: differential NCD burden (higher rates of diabetes, obesity, hypertension, and cardiovascular disease among Saudi nationals than the predominantly young expatriate workforce^18,21,22^); documented structural and access patterns favoring hospital-based over primary care utilization^26,27^); and broader systemic differences in healthcare access patterns between Saudi nationals and expatriates. Alattas and colleagues’ 2024 household survey confirmed that these patterns are further complicated by regional, wealth, and educational inequities in NCD service access.^29^ The narrow confidence intervals around the Saudi nationality ORs reflect the statistical power of 82,160 patients rather than conceptual precision: this single binary variable captures multiple simultaneously operating structural processes — NCD burden, care-seeking preferences, and differential access patterns — that would ideally be measured and modelled separately.

### Hospital Variation and Intervention Implications

The significant hospital × utilizer status interactions across all three ANOVA models and the between-facility range in FU intra-hospital prevalence (1.73% at TH to 3.54% at KAAH) confirm that the frequent utilization burden is not uniformly distributed and cannot be addressed by a system-wide generic response. KAAH’s elevated burden relative to EJH and TH is most plausibly associated with its tertiary role generating referral-driven clinical complexity across all patient groups — as evidenced by Non-FU mean bed days of 7.39 at KAAH versus 5.48 at EJH and 3.44 at TH. This implies that KAAH’s FU burden may be less reducible through primary care strengthening alone and may require a different intervention model centered on post-discharge coordination and specialist chronic disease management rather than primary care diversion.

The variation across hospitals reinforces a broader point about the design of interventions. Reducing frequent inpatient utilization requires not just identifying frequent utilizers but designing integrated clinical pathways that connect primary care and community health services to hospital care.^38,41^ Although most intervention effectiveness evidence derives from emergency department frequent user programmes,^11,12,13,39^ the case management and pathway principles are directly transferable to the inpatient context. For compound utilizers and PU-only patients, whose hospital dependence spans multiple years, a pathway that begins at the primary care center — with proactive identification, structured chronic disease management, and a designated care coordinator — and connects to hospital specialty services through shared protocols is the model most likely to reduce preventable admission and improve continuity.

### Policy Implications

Three actions are immediately implementable using existing administrative infrastructure. First, multi-definition surveillance dashboards should be integrated into hospital information systems to enable real-time identification of patients approaching the FU, PU, and YU thresholds — shifting from retrospective audit to prospective case management. Second, PU-only patients should be actively flagged in EHR systems through cross-year look-back queries; these 143 patients are structurally invisible under standard monitoring yet carry a median of 10.27 bed days — nearly four times the standard patient burden. Third, the concentration of high utilization in patients with a median age of 24 years, combined with the adolescent peak in persistent-utilization odds, signals an urgent need for dedicated young chronic disease pathways with structured pediatric-to-adult care transitions — a gap that administrative data can identify but only integrated clinical systems can close.

### Strengths and Limitations

This study’s principal strengths are its population-based scale (82,160 patients, 100,685 valid visits), multi-hospital design, three-year observation window, three-definition analytical framework, and transparent eight-stage data harmonization protocol. The identification of the composite key requirement — file numbers not being unique across hospitals — is an important data governance contribution for future multi-hospital Saudi administrative data research.

Five limitations require acknowledgement. First, only inpatient admissions at three Jeddah government hospitals are captured; primary care, outpatient, and emergency department utilization are absent, substantially underestimating total healthcare burden and system-wide FU prevalence. Second, ICD-10 diagnostic codes were used for encounter deduplication but not retained for patient-level analysis, precluding Charlson-based comorbidity adjustment^43^ and direct confirmation of the NCD and chronic disease mechanisms inferred throughout. This is the most significant limitation: the clinical framing rests on demographic inference and literature-based reasoning rather than direct diagnostic evidence. Third, bed-day utilization estimates are a proxy that does not account for variation in actual cost by specialty, severity, or ward type. Fourth, cross-sectional design precludes causal inference. Fifth, patients distributing admissions across multiple sectors will have their utilization underestimated.

## Conclusions

Approximately one in thirty-four inpatients across this three-hospital Jeddah system meets the rolling-window FU criterion. The three-definition framework reveals clinically and demographically distinct utilization phenotypes that no single threshold can identify: 143 PU-only patients with multi-year hospital dependence entirely invisible under annual-window definitions; 177 compound utilizers with extraordinary resource intensity and unexpectedly young age; and consistent gradients of increasing Saudi nationality concentration and female predominance as utilization complexity increases. Saudi nationality is the strongest and most consistent independent predictor across all definitions, findings consistent with interacting structural access, NCD burden, and healthcare-seeking mechanisms.

For health system planners implementing the Vision 2030 Health Sector Transformation Programme,25,40 these findings support five interconnected priorities. First, establish routine compound utilizer surveillance using existing administrative systems to enable early case management referral before patients reach the FU threshold.^10,38^ Second, design and implement integrated clinical pathways^41,42^ that formally connect primary care, community health services, and hospital specialty care for patients with chronic conditions driving frequent admission — pathways in which accountability for quality and resource utilization is shared across all levels of the health system rather than borne by the hospital alone. Third, develop intensive case management pathways targeting compound utilizers — the 177 patients satisfying all three utilization criteria simultaneously — who despite a median age of only 24 years consume a median of 33.44 bed days, representing the highest-burden and most identifiable priority group for intervention. Fourth, commission a follow-up study linking inpatient administrative data with primary care records, emergency department data, and ICD-10 diagnostic codes to enable disease-specific profiling, pathway evaluation, and assessment of whether Vision 2030-aligned investments measurably reduce frequent hospitalization over time. Fifth, establish structured pediatric-to-adult care transition protocols for adolescent patients with persistent multi-year hospital dependence. In both PU and YU Model 2, the Adolescent group (13–17 years) demonstrated the highest adjusted odds for persistent utilization relative to all older age groups, suggesting that sustained hospital engagement frequently originates in adolescence and continues into early adulthood without a coordinated transition in care. Linking pediatric and adult care within the same administrative system would enable prospective identification of this group before the transition gap widens.

## Declarations

### Abbreviations

ANOVA: Analysis of variance; CI: Confidence interval; EHR: Electronic health record; EJH: East Jeddah Hospital; FU: Frequent Utilizer; GCC: Gulf Cooperation Council; ICD-10: International Classification of Diseases, Tenth Revision; IQR: Interquartile range; IRB: Institutional Review Board; KAAH: King Abdul-Aziz Hospital; LOS: Length of stay; NCD: Non-communicable disease; OR: Odds ratio; PU: Persistent Utilizer; SD: Standard deviation; STROBE: Strengthening the Reporting of Observational Studies in Epidemiology; TH: Thagher Hospital; YU: Yearly Utilizer.

### Ethics approval and consent to participate

This study was approved by the Institutional Review Board of Jeddah Health Affairs (KACST registration number KSA: H-02-J-002; IRB approval number A02254; initial approval 16 July 2025; final approval 18 May 2026). As the study used routinely collected, de-identified administrative health record data involving no direct patient contact or intervention, the requirement for individual informed consent to participate was not applicable and was waived by the Institutional Review Board — Jeddah in accordance with KACST Good Clinical Practice regulations.

### Consent for publication

Not applicable. This study reports only aggregate statistical findings; no individual patient data, images, or other identifiable material are presented.

### Availability of data and materials

Aggregate statistical results are available upon reasonable request to the corresponding author. Individual-level patient data cannot be shared due to institutional confidentiality requirements.

### Competing interests

The authors declare no competing interests.

### Funding

No external funding was received for this study.

### Authors’ contributions

SB and RAR conceived and designed the study. AJA and ZA led the acquisition, cleaning, and analysis of the data. All authors participated in the interpretation of the results, reviewed the manuscript, and approved the final version.

## Acknowledgements

The authors thank the IT departments and management of East Jeddah Hospital, King Abdul-Aziz Hospital, and Thagher Hospital for their support in data provision, and acknowledge the Jeddah First Health Cluster for their institutional support throughout this study.

## Notes

### Competing Interest Statement

The authors have declared no competing interest.

### Author Declarations

The Institutional Review Board of Jeddah Health Affairs, Ministry of Health, Saudi Arabia gave ethical approval for this work (KACST registration number KSA: H-02-J-002; IRB approval number A02254). As the study used routinely collected, de-identified administrative health record data involving no direct patient contact or intervention, the requirement for individual informed consent was waived by the same board in accordance with KACST Good Clinical Practice regulations.

